# Clinician Counseling and Patient Storage and Disposal Practices for Opioids Prescribed for Acute Pain: A Multicenter Prospective Cohort Study

**DOI:** 10.64898/2026.11.04.26367278

**Authors:** Erfan Taherifard, Mitra Ahadpour, Fernanda Bellolio, Nancy Chang, Summer Allen, Marie-Carmelle Elie-Turenne, W. Jonathan Fillmore, Gregg H. Gilbert, Rebekah Heckmann, Patricia Koussis, Christine Lee, Heather Lipkind, Celeste Mallama, Tamra E. Meyer, Teryl K. Nuckols, Jessica D. Ritchie, Michael A. Pacanowski, David B. Page, Mat Soukup, Stephen Tamang, Sam Torbati, Doug Wallace, Lauren Walter, Yueqin Zhao, Nilay D. Shah, Joseph S. Ross, Molly Moore Jeffery

## Abstract

**BACKGROUND:** Opioids prescribed for acute pain frequently remain unused, creating opportunities for unintended use, accidental ingestion, or diversion. Little is known about counseling patients receive from prescribers or pharmacists on storage and disposal of opioids.

**OBJECTIVE:** To characterize clinician counseling on opioid disposal and patient-reported storage, use, and disposal practices among opioid-naïve patients prescribed opioids for acute pain.

**DESIGN:** Multicenter prospective cohort study with participant recruitment between September 2020 and March 2023.

**PARTICIPANTS:** Opioid-naïve patients prescribed opioids for acute pain in emergency departments, primary care or urgent care clinics, inpatient settings, and dental practices.

**MAIN MEASURES:** Patient-reported receipt of counseling on the importance of opioid disposal and appropriate disposal methods; opioid storage practices; use and disposition of prescribed opioids; disposal methods; and reasons for keeping or disposing of leftover opioids.

**KEY RESULTS:** Among 1708 participants, the median age was 38 years (IQR, 28-52); most were recruited from emergency departments (n=915; 53.6%) and primary care or urgent care clinics (n=307; 18.0%). Among 1233 participants who answered the question on counseling about the importance of opioid disposal, 811 (65.8%) reported receiving no counseling. Among 1220 participants who answered the question on counseling about appropriate disposal methods, 832 (68.2%) reported receiving no counseling. Among 812 participants who answered the question about storing opioids in a locked location, 610 (75.1%) reported never doing so. Among 929 participants providing information on opioid leftovers and disposition, 618 (66.5%) did not use all prescribed opioids. Among these 618 participants, 258 (42.8%) kept leftover opioids and 345 (57.2%) disposed of them. Among 256 participants who reported reasons for keeping leftover opioids, 159 (62.1%) wanted medication available for the same condition, 70 (27.3%) wanted medication available for a different condition, and 36 (14.1%) did not know how to properly dispose of the medication.

**CONCLUSIONS:** Among opioid-naïve patients prescribed opioids for acute pain, counseling about opioid disposal was uncommon, secure storage was infrequent, and leftover opioids were frequent, with many patients keeping leftover opioids for possible future use. These findings suggest opportunities to improve counseling on opioid storage and disposal and encourage safe disposal of unused opioids after acute pain treatment.

## INTRODUCTION

Opioid analgesics are frequently prescribed for the management of acute pain across diverse clinical settings.^1^ Although opioids can provide effective pain relief when prescribed and used appropriately, their use is associated with serious harms, including adverse effects, tolerance, dependence, misuse, overdose, and death.^2–5^ Despite the dominant role of illicitly made synthetic opioids in overdose deaths, prescription opioid-related harms remain a major public health concern in the United States; in 2025, over 7,000 overdose deaths involved prescription opioids,^6^ and 6.9 million people aged 12 years or older, including approximately 402,000 adolescents aged 12 to 17 years, reported misusing prescription opioids.^7^ Among people who misused prescription pain relievers, many obtained them from friends or relatives, including receiving them for free, buying them, or taking them without asking, underscoring the importance of reducing unused prescription opioids in the home.^8,9^

Unused opioids after treatment for acute pain may contribute to the continued availability of prescription opioids in the home, where they may later be used without medical supervision, shared with or taken by others, or accidentally ingested. Unintentional opioid exposure is particularly concerning among young children, with 2,746 cases involving children aged one month to six years reported to US poison centers in 2023.^10^ Prior studies have shown that patients prescribed opioids for postoperative or procedure-related pain frequently receive more tablets than they use, with a substantial proportion of prescribed opioids remaining unused. ^11–13^ However, much of the existing evidence has focused on postoperative populations, often with limited sample sizes, and less is known about storage and disposal practices among patients prescribed opioids for acute pain across broader clinical settings. Safe storage and timely disposal of unused opioids are therefore important strategies to reduce opioid availability in the home after acute pain treatment. The US Food and Drug Administration recommends medication take-back programs, including mail-back envelopes,^14^ as the preferred method for disposing of most unused medications, with flushing recommended for most opioid medicines when take-back options are not readily available.^15^

Counseling by prescribers and pharmacists may help patients understand the importance of opioid disposal and appropriate disposal methods,^16^ but evidence remains limited on counseling, storage, and disposal practices among patients prescribed opioids for acute pain across clinical settings. In this study, we examined opioid-naïve patients enrolled in the Acute Pain Pathways (APP) study^17,18^ who were prescribed opioid analgesics for acute pain to characterize opioid disposal counseling, patient-reported opioid storage practices, use and disposition of prescribed opioids, disposal methods used, and reasons for keeping or disposing of leftover opioids.

## METHODS

The APP study (NCT04509115) was a multicenter prospective cohort study that enrolled opioid-naïve patients between September 2020 and March 2023 who were prescribed opioid analgesics for the management of acute pain across multiple clinical settings, including emergency departments, primary care and urgent care clinics, inpatient settings, and dental practices. Patients were recruited from Cedars-Sinai Medical Center (California), Mayo Clinic (Minnesota, Florida, Wisconsin), Monument Health (South Dakota), University of Alabama at Birmingham (Alabama), Yale-New Haven Health (Connecticut), and community dental practices affiliated with the National Dental Practice-Based Research Network in several Southeastern states. Detailed eligibility criteria, recruitment procedures, and data collection methods for the APP study have been published previously.^17,18^ The study protocol was approved by the institutional review board at each participating site, and all participants provided written informed consent prior to enrollment. This study was reported in accordance with the STROBE guidelines.

### Study Population

Eligible participants in the APP study included adults aged 18 years or older at all participating sites and adolescents aged 15 years or older undergoing impacted third molar extraction at the Mayo Clinic in Rochester, Minnesota. Participants had acute pain of less than eight weeks’ duration at the time of enrollment, reported no opioid use in the preceding six months, and were offered an outpatient prescription for an opioid analgesic for pain management, including participants discharged from inpatient hospitalizations with an opioid prescription. Individuals with cancer-associated pain, end-of-life pain, or other pain conditions likely to progress to chronic pain were excluded, as were those who declined participation.

### Data Collection

Participants completed baseline and follow-up surveys through the Hugo Health digital platform, accessible through personal smartphones or other internet-enabled devices. Baseline surveys collected information on sociodemographic characteristics, clinical history, pain characteristics, treatments received, and social determinants of health. Follow-up surveys captured daily pain intensity and opioid and nonopioid use, as well as weekly assessments of side effects and patient-reported outcomes.

Opioid storage practices were assessed through weekly surveys, using the first available weekly response for each participant. When weekly survey data were unavailable, the end-of-study survey responses summarizing storage practices over the whole follow-up period were used. Outcomes related to use of all prescribed opioids, keeping or disposing of leftover opioids, disposal methods, and reasons for keeping or disposing of leftover opioids were assessed through the survey administered 2 weeks after cessation of opioid use or the end-of-study survey; the last available response was used to capture the final disposition of opioid medications. Receipt of counseling on the importance of opioid disposal and appropriate disposal methods was assessed through the week 1 survey, administered 1 week after enrollment. Instructions received about when to start and stop taking opioids were assessed once at enrollment.

### Outcomes

The counseling outcomes were patient-reported receipt of counseling on the importance of opioid disposal and receipt of counseling on appropriate disposal methods. For each counseling outcome, participants reported whether counseling was received and whether it was provided only by a pharmacist, only by a prescriber, or by both a pharmacist and a prescriber. Receipt of counseling was defined as reporting counseling by a pharmacist, prescriber, or both. Among participants who reported receipt of counseling on appropriate disposal methods, we characterized the specific disposal methods recommended by the clinician, including return to a medication take-back location, flushing down the toilet, mixing with an undesirable substance, and use of a disposal kit. Participants could report more than one recommended disposal method. We also characterized patient-reported instructions on when to take and stop taking opioids.

Storage-related outcomes included patient-reported frequency of storing opioids in a latched location and in a locked location, each assessed using response options of never, almost never, sometimes, almost always, and always. We also defined a composite storage outcome of storing opioids in a latched or locked location as reporting almost always or always for either storage in a latched location or storage in a locked location. We characterized the type of storage container used, including original prescription bottle, daily or weekly pill organizer, single-compartment container, or other container.

Disposal-related outcomes included whether participants used all prescribed opioids, whether participants kept leftover opioids among those who did not use all prescribed opioids, and disposal methods used among participants who disposed of leftover opioids. We also characterized patient-reported reasons for disposing of or keeping leftover opioids among participants who disposed of or kept them, respectively. Participants could report more than one disposal method or reason.

### Covariates

Covariates included age group (≤21, 22-34, 35-44, 45-54, 55-64, and ≥65 years), presence of children in the household, race/ethnicity (Asian, American Indian or Alaska Native, Black or African American, Hispanic or Latino, Native Hawaiian or other Pacific Islander, White, other, or unknown), residential setting (urban, rural, or unknown), education level (less than high school, high school diploma or General

Educational Development, more than high school, or unknown), clinical setting of care (emergency departments, primary care or urgent care clinics, inpatient settings, or dental practices), and pain source (dental, obstetric, surgical or procedural, trauma or injury, musculoskeletal, abdominal, renal colic/kidney stone, skin or soft tissue, low back pain, other, or unknown).

### Statistical Analysis

Data management and statistical analyses were performed using Stata version 18 (StataCorp LLC, College Station, TX, USA). Categorical variables were summarized using counts and percentages, and continuous variables using medians and interquartile ranges (IQRs). Survey response timing was summarized as the elapsed number of days from prescription offer to the survey response. We used χ² tests to compare categorical outcomes across groups.

To estimate adjusted associations, we fit multivariable logistic regression models with robust standard errors and calculated risk ratios (RRs) as ratios of predictive margins. Models for receipt of counseling on the importance of opioid disposal, receipt of counseling on appropriate disposal methods, and storing opioids in a latched or locked location included age group, clinical setting, pain source, and presence of children in the household. The model for keeping leftover opioids was restricted to participants who did not use all prescribed opioids and included age group, clinical setting, and presence of children in the household; pain source was not included because of sparse cells and perfect prediction. Due to missing data in the variable for whether people had leftover opioids, we used a multiple imputation analysis to assess the potential impact of this missing data; the analysis is described in the Supplement. A 2-sided P value of less than 0.05 was considered statistically significant. This analysis was conducted as part of the larger APP study, which was not specifically designed to detect differences in opioid counseling, storage, and disposal outcomes across participant groups. Therefore, these analyses were considered exploratory.

## RESULTS

### Study Population

Among 1708 participants prescribed opioids for acute pain, 915 (53.6%) were recruited from emergency departments, 307 (18.0%) from primary care or urgent care clinics, 263 (15.4%) from dental practices, and 223 (13.1%) from inpatient settings (**Table 1**). Most participants were aged 22 to 34 years (n=551; 32.3%) or 35 to 44 years (n=368; 21.6%), with a median age of 38 years (IQR, 28-52). Most participants were White (n=1093; 64.0%) and had more than a high school education (n=906; 67.0%). The most common pain sources were dental pain (n=302; 17.7%), trauma or injury (n=302; 17.7%), and obstetric pain (n=176; 10.3%). Among 1144 participants (67.0%) with available household information, 814 (71.2%) had children in the household. Participant characteristics differed significantly across clinical settings, including age group, presence of children in the household, race/ethnicity, residential setting, educational level, and pain source (**Supplementary Table 1**). Data on opioid disposal were available for 929 participants (54.4%). Compared with participants without available data, those with available data differed significantly by age group, clinical setting, race/ethnicity, educational level, and pain source.

**Table 1.** Characteristics of Opioid-Naïve Participants Prescribed Opioids for Acute Pain, Overall and by Availability of Opioid Use Data.

| Characteristic | Total,<br>No. (%) | Participants with data on opioid leftovers,<br>No. (%) | Participants without data on opioid leftovers,<br>No. (%) | P value |
|---|---|---|---|---|
| All participants | 1708 (100) | 929 (100) | 779 (100) |  |
| Age group |  |  |  |  |
| 21 and under | 170 (10.0) | 95 (10.2) | 75 (9.6) | 0.007 |
| 22-34 | 551 (32.3) | 266 (28.6) | 285 (36.6) |  |
| 35-44 | 368 (21.6) | 206 (22.2) | 162 (20.8) |  |
| 45-54 | 267 (15.6) | 146 (15.7) | 121 (15.5) |  |
| 55-64 | 216 (12.7) | 135 (14.5) | 81 (10.4) |  |
| 65 and older | 136 (8.0) | 81 (8.7) | 55 (7.1) |  |
| Clinical setting |  |  |  |  |
| Emergency departments | 915 (53.6) | 480 (51.7) | 435 (55.8) | <0.001 |
| Primary care or urgent care clinics | 307 (18.0) | 200 (21.5) | 107 (13.7) |  |
| Inpatient settings | 223 (13.1) | 104 (11.2) | 119 (15.3) |  |
| Dental practice | 263 (15.4) | 145 (15.6) | 118 (15.1) |  |
| Children in household |  |  |  |  |
| Yes | 814 (71.2) | 539 (69.7) | 275 (74.1) | 0.12 |
| No | 330 (28.8) | 234 (30.3) | 96 (25.9) |  |
| Did not respond to survey | 564 | 156 | 408 | — |
| Race/ethnicity |  |  |  |  |
| White | 1093 (64.0) | 623 (67.1) | 470 (60.3) | 0.012 |
| Black or African American | 298 (17.5) | 142 (15.3) | 156 (20.0) |  |
| Hispanic or Latino | 155 (9.1) | 69 (7.4) | 86 (11.0) |  |
| American Indian or Alaska Native | 64 (3.8) | 36 (3.9) | 28 (3.6) |  |
| Asian | 43 (2.5) | 25 (2.7) | 18 (2.3) |  |
| Native Hawaiian or other Pacific Islander | 7 (0.4) | 5 (0.5) | 2 (0.3) |  |
| Other | 48 (2.8) | 29 (3.1) | 19 (2.4) |  |
| Residential setting |  |  |  |  |
| Urban | 647 (80.8) | 396 (78.9) | 251 (83.9) | <0.08 |
| Rural | 154 (19.2) | 106 (21.1) | 48 (16.1) |  |
| Did not answer | 353 | 79 | 274 | — |
| Did not respond to survey | 554 | 348 | 206 | — |
| Educational level |  |  |  |  |
| Less than high school | 104 (7.7) | 77 (9.0) | 27 (5.5) | <0.001 |
| High school diploma or GED | 342 (25.3) | 190 (22.2) | 152 (30.7) |  |
| More than high school | 906 (67.0) | 590 (68.8) | 316 (63.8) |  |
| Did not answer | 72 | 25 | 47 | — |
| Did not respond to survey | 284 | 47 | 237 | — |
| Pain source |  |  |  |  |
| Dental | 302 (17.7) | 165 (17.8) | 137 (17.6) | <0.001 |
| Obstetric | 176 (10.3) | 81 (8.7) | 95 (12.2) |  |
| Surgical or procedural | 65 (3.8) | 34 (3.7) | 31 (4.0) |  |
| Trauma or injury | 302 (17.7) | 184 (19.8) | 118 (15.1) |  |
| Musculoskeletal | 131 (7.7) | 84 (9.0) | 47 (6.0) |  |
| Abdominal | 98 (5.7) | 68 (7.3) | 30 (3.9) |  |
| Kidney stone | 88 (5.2) | 61 (6.6) | 27 (3.5) |  |
| Skin or soft tissue | 85 (5.0) | 48 (5.2) | 37 (4.7) |  |
| Low back pain | 100 (5.9) | 70 (7.5) | 30 (3.9) |  |
| Other | 86 (5.0) | 51 (5.5) | 35 (4.5) |  |
| Unknown in emergency department | 233 (13.6) | 66 (7.1) | 167 (21.4) |  |
| Unknown in primary care or urgent care clinics | 42 (2.5) | 17 (1.8) | 25 (3.2) |  |

**Note:** Percentages are calculated among participants with nonmissing responses for each characteristic. “Did not answer” indicates participants who responded to some questions on the relevant survey but did not respond to that item. “Did not respond to survey” indicates participants who did not complete the relevant survey.
**Abbreviation:** GED, General Educational Development.

### Counseling and Instructions on Opioid Use and Disposal

Among 1233 participants (72.2%) who answered the question on counseling about the importance of opioid disposal, 811 (65.8%) did not receive counseling, 231 (18.7%) received counseling only from a pharmacist, 140 (11.4%) only from a prescriber, and 51 (4.1%) from both (**Table 2**). Among 1220 participants (71.4%) who answered the question on counseling about appropriate disposal methods, 832 (68.2%) did not receive counseling, 217 (17.8%) received counseling only from a pharmacist, 119 (9.8%) only from a prescriber, and 52 (4.3%) from both. Among 388 participants who received counseling on appropriate disposal methods, 324 (83.5%) provided information on the specific disposal methods recommended by the clinician; recommended methods included return to a take-back location (n=221; 68.2%), flushing down the toilet (n=73; 22.5%), mixing with an undesirable substance (n=28; 8.6%), and use of a disposal kit (n=25; 7.7%).

**Table 2.** Clinician Counseling on Importance of Opioid Disposal and Appropriate Disposal Methods by Clinical Setting.

|  | Total,<br>No. (%) | Emergency departments,<br>No. (%) | Primary care or urgent care clinics,<br>No. (%) | Inpatient settings,<br>No. (%) | Dental practice,<br>No. (%) | P Value |
|---|---|---|---|---|---|---|
| All participants | 1708 (100) | 915 (53.6) | 307 (18.0) | 223 (13.1) | 263 (15.4) | — |
| Receipt of counseling on importance of opioid disposal |  |  |  |  |  |  |
| Not received | 811 (65.8) | 428 (65.5) | 173 (67.8) | 89 (64.0) | 121 (65.1) | 0.45 |
| Provided only by pharmacist | 231 (18.7) | 119 (18.2) | 44 (17.3) | 23 (16.6) | 45 (24.2) |  |
| Provided only by prescriber | 140 (11.4) | 77 (11.8) | 27 (10.6) | 21 (15.1) | 15 (8.1) |  |
| Provided by both | 51 (4.1) | 29 (4.4) | 11 (4.3) | 6 (4.3) | 5 (2.7) |  |
| Did not answer | 475 | 262 | 52 | 84 | 77 | — |
| Receipt of counseling on appropriate disposal methods |  |  |  |  |  |  |
| Not received | 832 (68.2) | 447 (68.9) | 177 (69.7) | 85 (64.9) | 123 (66.1) | 0.01 |
| Provided only by pharmacist | 217 (17.8) | 113 (17.4) | 42 (16.5) | 16 (12.2) | 46 (24.7) |  |
| Provided only by prescriber | 119 (9.8) | 59 (9.1) | 24 (9.4) | 24 (18.3) | 12 (6.5) |  |
| Provided by both | 52 (4.3) | 30 (4.6) | 11 (4.3) | 6 (4.6) | 5 (2.7) |  |
| Did not answer | 488 | 266 | 53 | 92 | 77 | — |
| Disposal methods recommended by the clinician (n=388) |  |  |  |  |  |  |
| Return to take-back location |  |  |  |  |  |  |
| Yes | 221 (68.2) | 117 (68.4) | 47 (72.3) | 26 (68.4) | 31 (62.0) | 0.71 |
| No | 103 (31.8) | 54 (31.6) | 18 (27.7) | 12 (31.6) | 19 (38.0) |  |
| Did not answer | 64 | 31 | 12 | 8 | 13 | — |
| Flush down the toilet |  |  |  |  |  |  |
| Yes | 73 (22.5) | 40 (23.4) | 11 (16.9) | 12 (31.6) | 10 (20.0) | 0.36 |
| No | 251 (77.5) | 131 (76.6) | 54 (83.1) | 26 (68.4) | 40 (80.0) |  |
| Did not answer | 64 | 31 | 12 | 8 | 13 | — |
| Mix with an undesirable substance |  |  |  |  |  |  |
| Yes | 28 (8.6) | 14 (7.5) | 4 (5.5) | 3 (6.5) | 7 (14.0) | 0.49 |
| No | 296 (91.4) | 157 (92.5) | 61 (94.5) | 35 (93.5) | 43 (86.0) |  |
| Did not answer | 64 | 31 | 12 | 8 | 13 | — |
| Use of a disposal kit |  |  |  |  |  |  |
| Yes | 25 (7.7) | 13 (7.1) | 9 (13.8) | 0 (0.0) | 3 (6.0) | 0.08 |
| No | 299 (92.3) | 158 (92.9) | 56 (86.2) | 38 (100.0) | 47 (94.0) |  |
| Did not answer | 64 | 31 | 12 | 8 | 13 | — |

**Note:** Percentages are calculated among participants with nonmissing responses for each item. “Did not answer” indicates participants who reported receiving counseling on appropriate disposal methods but did not provide a response for that specific recommended method. Participants may have reported being recommended more than one disposal method.

Among 1328 participants (77.8%) who provided information on instructions received about when to take opioids, 522 (39.3%) reported being instructed to take opioids only when pain was very bad, 317 (23.9%) at the first sign of pain, and 291 (21.9%) to try other medications before taking opioids. Overall, 101 (7.6%) reported receiving no instructions on when to take opioids. Among 1302 participants (76.2%) who provided information on instructions received about when to stop taking opioids, 417 (32.0%) reported being instructed to continue taking opioids as long as they had pain, 289 (22.2%) until the pills ran out, and 177 (13.6%) after a specified number of days. Overall, 456 (35.0%) reported receiving no instructions on when to stop taking opioids. Instructions differed across clinical settings, with participants recruited from inpatient settings most frequently reporting no instructions on when to take opioids (17.8%) and participants recruited from dental practices most frequently reporting instructions to continue taking opioids as long as pain persisted (41.5%) (**Supplementary Table 2**).

In the adjusted model, receipt of counseling on the importance of opioid disposal and counseling on appropriate disposal methods did not differ significantly by age group, clinical setting, or presence of children in the household (**Supplementary Table 3**).

### Storage Practices

Among 1261 participants (73.8%) with available opioid storage survey responses, 1163 (92.2%) responses were obtained from weekly surveys and 98 (7.8%) from end-of-study surveys; responses were recorded at a median of 16 days after prescription offer (IQR, 14-22 days). Among 877 participants who answered the question about storing opioids in a latched location, 352 (40.1%) reported never doing so, while 378 (43.1%) reported always doing so. Among 812 participants who answered the question about storing opioids in a locked location, 610 (75.1%) reported never doing so, while 141 (17.4%) reported always doing so (**Table 3; Supplementary Table 4**). Among 473 participants who reported having children in the household and answered the question about locked opioid storage, 345 (72.9%) reported never storing opioids in a locked location. Most participants stored opioids in the original prescription bottle (n=797; 63.2%); few used a daily or weekly pill organizer (n=15; 1.2%) or single-compartment container (n=18; 1.4%).

**Table 3.** Storage and Disposal Practices of Opioid-Naïve Patients Prescribed Opioids for Acute Pain, Stratified by Age Group.

|  | Overall<br>No. (%) | Aged ≤21,<br>No. (%) | Aged 22-34,<br>No. (%) | Aged 35-44,<br>No. (%) | Aged 45-54,<br>No. (%) | Aged 55-64,<br>No. (%) | Aged ≥65,<br>No. (%) | P Value |
|---|---|---|---|---|---|---|---|---|
| All patients | 1708 (100) | 170 (100) | 551 (100) | 368 (100) | 267 (100) | 216 (100) | 136 (100) | — |
| Storing opioids in a latched location |  |  |  |  |  |  |  |  |
| Never | 352 (40.1) | 30 (40.0) | 93 (36.9) | 69 (35.6) | 58 (37.9) | 58 (45.3) | 44 (58.7) | 0.03 |
| Almost never | 57 (6.5) | 8 (10.7) | 14 (5.6) | 10 (5.2) | 8 (5.2) | 13 (10.2) | 4 (5.3) |  |
| Sometimes | 46 (5.3) | 7 (9.3) | 16 (6.3) | 9 (4.6) | 9 (5.9) | 2 (1.6) | 3 (4.0) |  |
| Almost always | 44 (5.0) | 5 (6.7) | 12 (4.8) | 11 (5.7) | 8 (5.2) | 8 (6.2) | 0 (0.0) |  |
| Always | 378 (43.1) | 25 (33.3) | 117 (46.4) | 95 (49.0) | 70 (45.8) | 47 (36.7) | 24 (32.0) |  |
| Did not use opioid | 384 | 47 | 124 | 85 | 54 | 46 | 28 | — |
| Did not answer | 447 | 48 | 175 | 89 | 60 | 42 | 33 | — |
| Storing opioids in a locked location |  |  |  |  |  |  |  |  |
| Never | 610 (75.1) | 58 (81.7) | 172 (72.9) | 124 (70.1) | 103 (71.5) | 96 (80.7) | 57 (87.7) | 0.004 |
| Almost never | 43 (5.3) | 6 (8.5) | 9 (3.8) | 8 (4.5) | 6 (4.2) | 10 (8.4) | 4 (6.2) |  |
| Sometimes | 18 (2.2) | 3 (4.2) | 5 (2.1) | 4 (2.3) | 3 (2.1) | 2 (1.7) | 1 (1.5) |  |
| Almost always | 0 (0.0) | 0 (0.0) | 0 (0.0) | 0 (0.0) | 0 (0.0) | 0 (0.0) | 0 (0.0) |  |
| Always | 141 (17.4) | 4 (5.6) | 50 (21.2) | 41 (23.2) | 32 (22.2) | 11 (9.2) | 3 (4.6) |  |
| Did not use opioid | 25 | 3 | 8 | 4 | 3 | 5 | 2 | — |
| Did not answer | 871 | 96 | 307 | 187 | 120 | 92 | 69 | — |
| Opioid storage location |  |  |  |  |  |  |  |  |
| Original prescription bottle |  |  |  |  |  |  |  |  |
| Yes | 797 (63.2) | 65 (53.3) | 227 (60.4) | 180 (64.5) | 142 (68.6) | 122 (70.1) | 61 (59.2) | 0.02 |
| No | 464 (36.8) | 57 (46.7) | 149 (39.6) | 99 (35.5) | 65 (31.4) | 52 (29.9) | 42 (40.8) |  |
| Did not answer | 447 | 48 | 175 | 89 | 60 | 42 | 33 | — |
| Daily or weekly pill organizer |  |  |  |  |  |  |  |  |
| Yes | 15 (1.2) | 0 (0.0) | 6 (1.6) | 2 (0.7) | 2 (1.0) | 2 (1.1) | 3 (2.9) | 0.39 |
| No | 1,246 (98.8) | 122 (100.0) | 370 (98.4) | 277 (99.3) | 205 (99.0) | 172 (98.9) | 100 (97.1) |  |
| Did not answer | 447 | 48 | 175 | 89 | 60 | 42 | 33 | — |
| Single-compartment container |  |  |  |  |  |  |  |  |
| Yes | 18 (1.4) | 2 (1.6) | 6 (1.6) | 4 (1.4) | 3 (1.4) | 3 (1.7) | 0 (0.0) | 0.89 |
| No | 1,243 (98.6) | 120 (98.4) | 370 (98.4) | 275 (98.6) | 204 (98.6) | 171 (98.3) | 103 (100.0) |  |
| Did not answer | 447 | 48 | 175 | 89 | 60 | 42 | 33 | — |
| Use of all prescribed opioids |  |  |  |  |  |  |  |  |
| Yes | 311 (33.5) | 22 (23.2) | 93 (35.0) | 73 (35.4) | 61 (41.8) | 41 (30.4) | 21 (25.9) | 0.03 |
| No | 618 (66.5) | 73 (76.8) | 173 (65.0) | 133 (64.6) | 85 (58.2) | 94 (69.6) | 60 (74.1) |  |
| Did not answer | 779 | 75 | 285 | 162 | 121 | 81 | 55 | — |
| Kept leftover opioids among those who did not use all opioids (n=618) |  |  |  |  |  |  |  |  |
| Yes | 258 (42.8) | 21 (30.0) | 51 (30.4) | 56 (42.7) | 48 (57.1) | 51 (54.8) | 31 (54.4) | <0.001 |
| No | 345 (57.2) | 49 (70.0) | 117 (69.6) | 75 (57.3) | 36 (42.9) | 42 (45.2) | 26 (45.6) |  |
| Did not answer | 15 | 3 | 5 | 2 | 1 | 1 | 3 | — |
| Disposal methods used among those who disposed of leftover opioids (n=345) |  |  |  |  |  |  |  |  |
| Return to take-back location |  |  |  |  |  |  |  |  |
| Yes | 114 (40.3) | 19 (46.3) | 41 (42.3) | 19 (33.3) | 9 (26.5) | 16 (47.1) | 10 (50.0) | 0.30 |
| No | 169 (59.7) | 22 (53.7) | 56 (57.7) | 38 (66.7) | 25 (73.5) | 18 (52.9) | 10 (50.0) |  |
| Did not answer | 62 | 8 | 20 | 18 | 2 | 8 | 6 | — |
| Flush down the toilet |  |  |  |  |  |  |  |  |
| Yes | 105 (37.1) | 12 (29.3) | 33 (34.0) | 26 (45.6) | 16 (47.1) | 11 (32.4) | 7 (35.0) | 0.41 |
| No | 178 (62.9) | 29 (70.7) | 64 (66.0) | 31 (54.4) | 18 (52.9) | 23 (67.6) | 13 (65.0) |  |
| Did not answer | 62 | 8 | 20 | 18 | 2 | 8 | 6 | — |
| Throw away in the trash |  |  |  |  |  |  |  |  |
| Yes | 67 (23.7) | 12 (29.3) | 23 (23.7) | 14 (24.6) | 9 (26.5) | 6 (17.6) | 3 (15.0) | 0.79 |
| No | 216 (76.3) | 29 (70.7) | 74 (76.3) | 43 (75.4) | 25 (73.5) | 28 (82.4) | 17 (85.0) |  |
| Did not answer | 62 | 8 | 20 | 18 | 2 | 8 | 6 | — |

**Note:** Percentages for storage-related items are calculated among participants with nonmissing responses who reported opioid use. “Did not answer” indicates participants who responded to some questions on the relevant survey but did not respond to that item. “Did not use opioid” indicates participants who reported no opioid use at the time of survey completion. Participants may have reported more than one disposal method.

In the adjusted model, participants with children in the household were more likely than those without children to store opioids in a latched or locked location (RR, 1.25; 95% CI, 1.05-1.48; P=0.01) (**Supplementary Table 5**). Clinical setting and age group were not significantly associated with storage in a latched or locked location.

### Prescribed Opioid Leftovers and Disposition

Among 929 participants (54.4%) with available responses on leftovers and disposition of prescribed opioids, 847 (91.2%) responses were obtained from end-of-study surveys and 82 (8.8%) from end-of-opioid-use surveys; responses were recorded at a median of 185 days after prescription offer (IQR, 181-241 days). Small but statistically significant differences in observed characteristics were found between respondents and participants with missing data (**Table 1**). Overall, 311 participants (33.5%) used all prescribed opioids and 618 (66.5%) did not (**Figure 1**). A multiple imputation analysis found a similar proportion of participants with leftover opioids (**Supplementary Table 6**). Among participants aged 21 years or younger, 73 of 95 (76.8%) did not use all prescribed opioids, compared with 545 of 834 (65.3%) participants aged 22 years or older (**Table 3**). Among 618 participants who did not use all prescribed opioids, 603 (97.6%) provided information on the disposition of leftover opioids; 258 (42.8%) kept the leftover opioids and 345 (57.2%) disposed of them. Among participants with leftover opioids who provided disposition information, 21 of 70 (30.0%) aged 21 years or younger kept them, compared with 237 of 533 (44.5%) aged 22 years or older. Among 283 participants who disposed of leftover opioids and answered questions about disposal methods, 114 (40.3%) returned them to a take-back location, 105 (37.1%) flushed them down the toilet, and 67 (23.7%) threw them in the trash.

**Figure 1.**
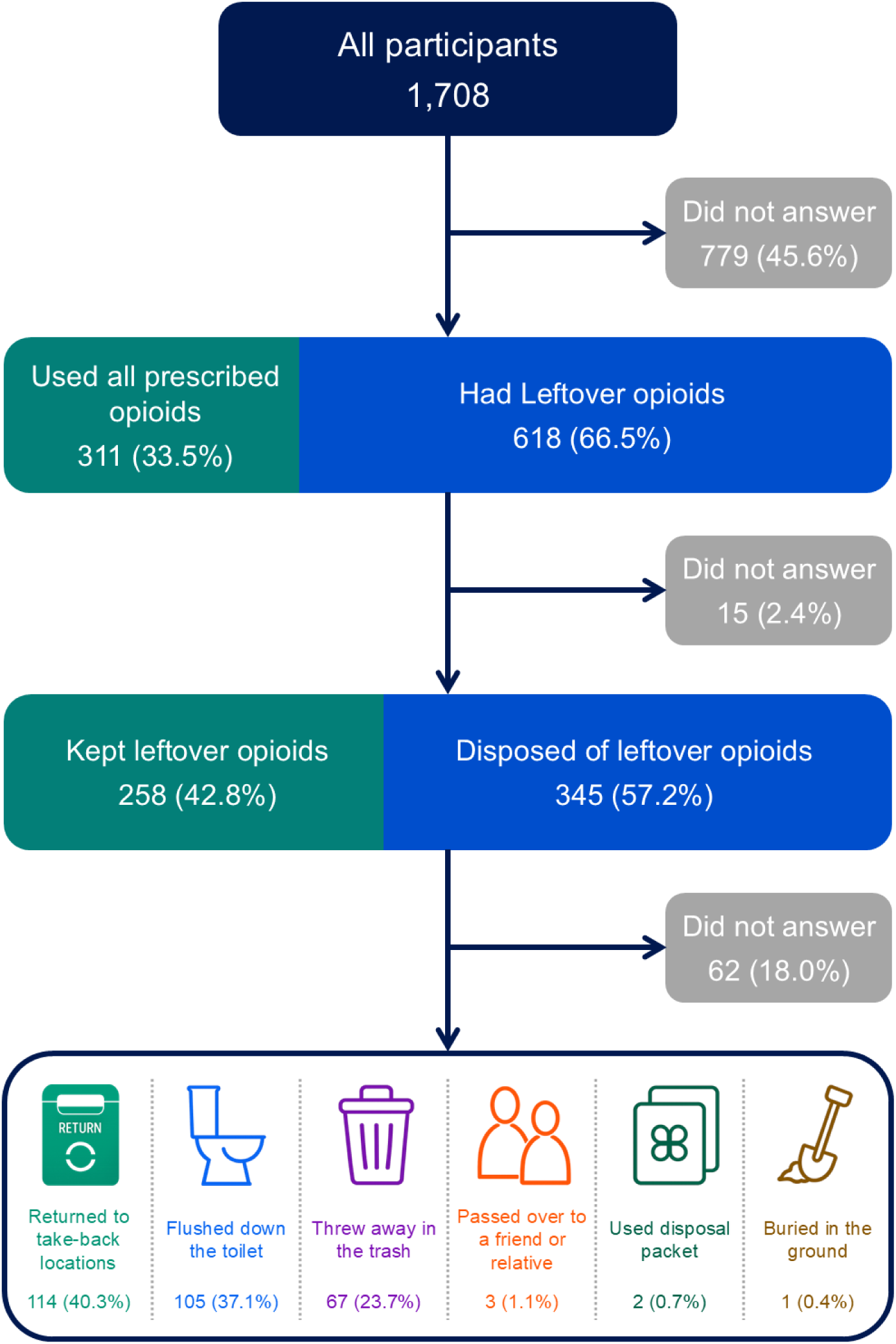
Disposition of Leftover Opioids Among Opioid-Naïve Participants Prescribed Opioids for Acute Pain. **Note:** The two horizontal boxes are stacked bars. The first stacked bar shows, among participants with available responses, the proportion who used all prescribed opioids and the proportion who had leftover opioids. The second stacked bar shows, among participants with leftover opioids and available responses, the proportion who kept leftover opioids and the proportion who disposed of leftover opioids.

Among participants who did not use all prescribed opioids, age group, clinical setting, and presence of children in the household were not significantly associated with keeping leftover opioids (**Supplementary Table 5**).

### Reasons for Disposal or Keeping Leftover Opioids

Among 345 participants who disposed of leftover opioids, 296 (85.8%) provided reasons for disposal; the most common reasons were routinely disposing of unused prescription medications (n=180; 60.8%), awareness of opioid-related risks (n=115; 38.9%), advice from a health care professional (n=48; 16.2%), and concern that a child might accidentally find or ingest the medication (n=43; 14.5%) (**Figure 2**).

**Figure 2.**
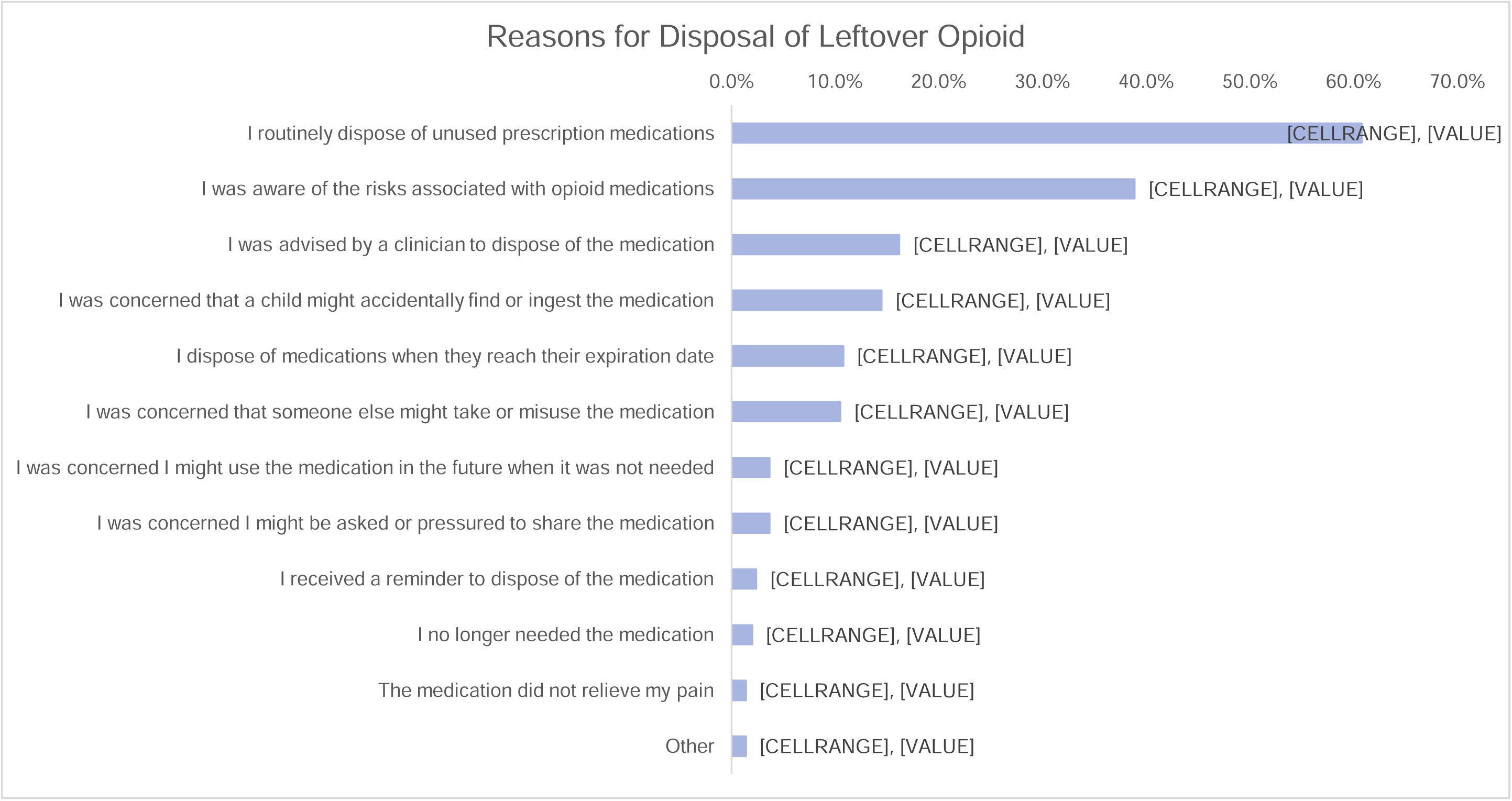
Reasons for Disposal of Leftover Opioids Among Opioid-Naïve Participants Prescribed Opioids for Acute Pain Who Disposed of the Leftover Opioids (n=296) **Note:** Participants could report more than one reason. Reasons are shown among participants who disposed of leftover opioids and provided responses to this question (n=296); 49 participants did not provide a response.

Among 258 participants who kept leftover opioids, 256 (99.2%) provided reasons for keeping them; the most common reasons were wanting medication available for the same condition (n=159; 62.1%), wanting medication available for a different condition (n=70; 27.3%), not knowing how to properly dispose of the medication (n=36; 14.1%), not wanting to discard medication because it still had value (n=27; 10.5%), and concerns about the environmental impact of disposal (n=26; 10.2%) (**Figure 3**).

**Figure 3.**
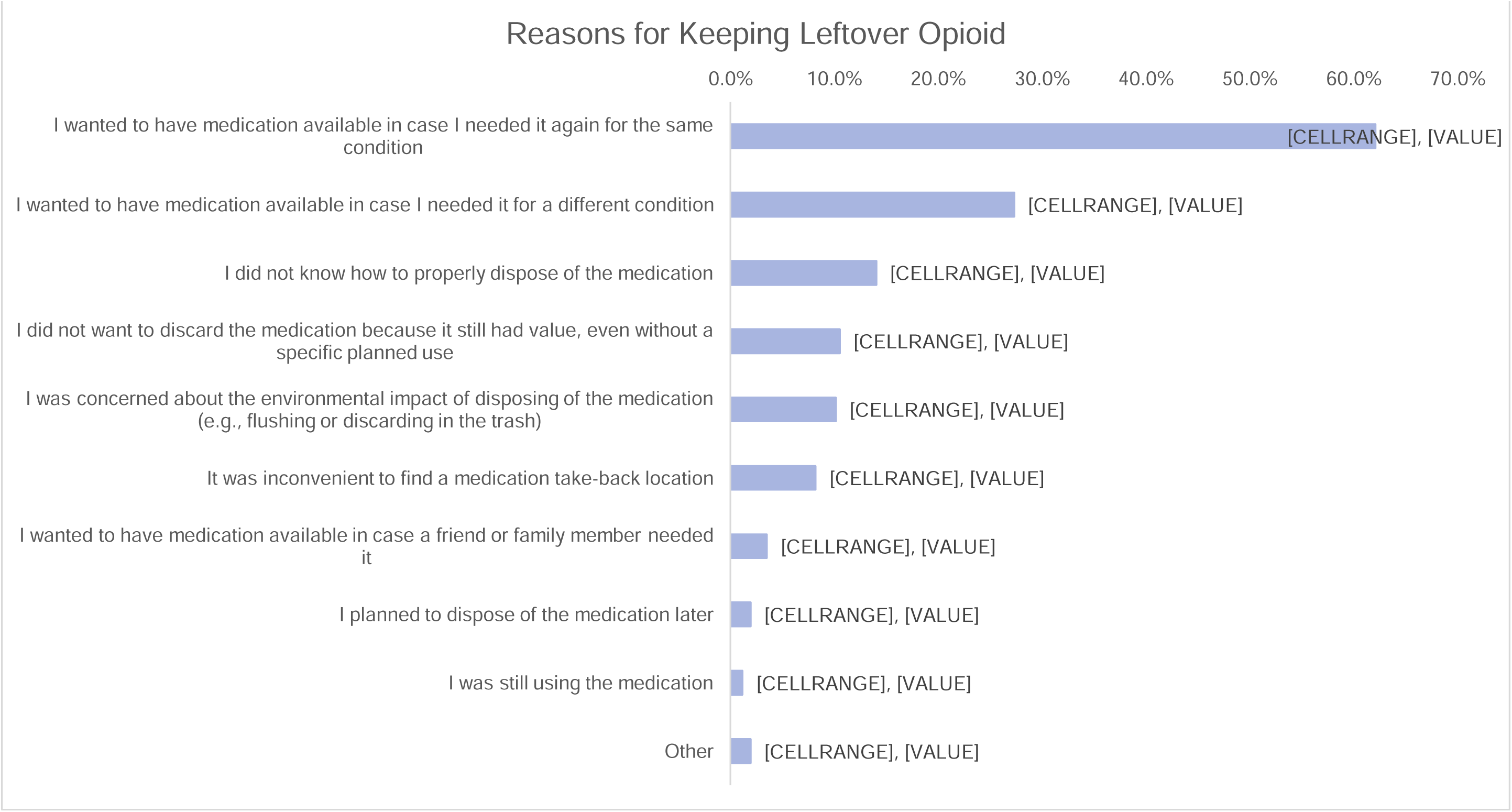
Reasons for Keeping Leftover Opioids Among Opioid-Naïve Participants Prescribed Opioids for Acute Pain Who Kept Leftover Opioids (n=256) **Note:** Participants could report more than one reason. Reasons are shown among participants who kept leftover opioids and provided responses to this question (n=256); 2 participants did not provide a response.

## DISCUSSION

In this multicenter prospective cohort study of opioid-naïve patients prescribed opioids for acute pain, approximately two-thirds of respondents reported no counseling on the importance of opioid disposal or appropriate disposal methods, with counseling rates similar across clinical settings and pain sources. Among participants who answered questions about locked storage, three-quarters reported never storing opioids in a locked location, although participants with children in the household were more likely to store opioids in a latched or locked location. Approximately two-thirds of participants with information on opioid leftovers and disposition did not use all prescribed opioids, and more than 40% of those with leftover opioids kept them, most commonly because they wanted to have the medication available for possible future use. Nearly 25% of those who disposed of leftover opioids did so in the trash, which is not recommended method for opioid disposal; instead, FDA recommends disposing of most opioids by flushing them down the toilet when take-back options or mail-back envelopes are not readily available.^15^ Together, these findings suggest opportunities to improve counseling, storage, and disposal practices and reduce the amount of unused opioids remaining after treatment for acute pain.

Counseling on the importance of opioid disposal and appropriate disposal methods was uncommon, with similar rates observed across clinical settings and pain sources. Among participants who received counseling, pharmacists were more commonly reported as the sole source of counseling than prescribers, highlighting the important role of pharmacists while also suggesting missed opportunities to reinforce counseling throughout the medication-use process. This finding is particularly relevant given current recommendations that prescribers and pharmacists counsel patients on secure storage and safe disposal of unused opioids.^1,19,20^ Previous studies have similarly found that counseling on safe opioid storage and disposal is not consistently provided to patients prescribed opioids.^21,22^ The similar counseling rates observed across clinical settings in our study, despite differences in clinical workflow and the amount and allocation of clinician-patient time across these settings,^23–25^ suggest that limited counseling may reflect a broader gap in counseling practices when opioids are prescribed for acute pain, rather than being concentrated within settings where opportunities for patient counseling may be more limited.

Most participants with available storage responses did not consistently store opioids in a secure location, with three-quarters of them reporting that they never stored their opioids in a locked location. Previous studies have similarly found that unused opioids are frequently stored in unsecured locations after treatment for acute pain, with systematic reviews reporting that approximately 70% or more of patients with leftover opioids did not store them in a locked location.^11,26^ Participants with children in the household in our study were more likely to store opioids in a latched or locked location than those without children, suggesting that concerns about children may influence storage practices. However, the risks of unsecured storage extend beyond children and include other family members and friends, who may access opioids kept in the home, including medications taken without asking.^7,27^ The presence of children was not associated with whether participants kept leftover opioids, suggesting that greater attention to secure storage may not necessarily translate into removing unused opioids from the home. These findings highlight the importance of counseling about both secure storage and disposal of unused opioids, regardless of whether children are present in the household.

Approximately two-thirds of participants who responded to questions on opioid leftovers and disposition did not use all of the opioids prescribed to them, and among those with leftover opioids, more than 40% kept them instead of disposing of them. Previous studies have similarly found that patients often keep unused opioids for possible future pain, suggesting that leftover medications may be viewed as useful for subsequent episodes of pain.^28,29^ In our study, having medication available for future use, either for the same condition or a different condition, was the most commonly reported reason for keeping leftover opioids, while not knowing how to properly dispose of them was reported much less frequently. This difference is important because providing instructions on how to dispose of unused opioids alone may have limited effects on reducing the amount of medication patients keep. Counseling may also need to address the potential risks of keeping and using leftover opioids for future episodes of pain without a new clinical assessment. In addition, prescribing quantities that more closely reflect patients’ expected needs may reduce the amount of unused opioids remaining after treatment for acute pain.

This study has several limitations. First, not all participants completed each follow-up survey or responded to each question, resulting in variation in the availability of data across study outcomes. In particular, information on opioid leftovers and disposition was available for approximately half of participants, and there were minor but statistically significant differences in observed characteristics between respondents and non-respondents, which may introduce selection bias. Second, counseling, instructions on opioid use, storage, opioid leftovers, and disposal practices were based on participant self-report and may be subject to recall or social desirability bias. Third, although we assessed whether participants reported receiving counseling on the importance of opioid disposal and appropriate disposal methods, we had limited information on the specific content, duration, or quality of that counseling beyond the disposal methods recommended. We therefore could not determine whether the counseling provided differed across clinical settings or whether differences in counseling content or quality were associated with subsequent opioid storage or disposal practices. Fourth, this analysis was conducted as part of the APP study, which was not specifically designed or powered to detect differences in opioid counseling, storage, and disposal outcomes across participant groups. Therefore, the analyses were exploratory. In addition, although we adjusted for several demographic and clinical characteristics, differences in patient populations across clinical settings, particularly the limited overlap in pain sources between dental practices and other settings, may have resulted in residual confounding and influenced the observed associations. We also did not adjust for the quantity of opioids prescribed, which may have influenced storage practices and leftover opioids. Lastly, although participants were recruited from multiple clinical settings and represented a range of acute pain conditions, all were opioid-naïve, which may limit the generalizability of these findings outside that population.

## CONCLUSIONS

In this multicenter prospective cohort study of opioid-naïve patients prescribed opioids for acute pain, we found that counseling about opioid disposal was uncommon, secure storage was infrequent, and leftover opioids were frequent, with many patients keeping unused opioids for possible future use. These findings suggest opportunities to improve counseling on secure opioid storage and disposal, including addressing the potential risks of keeping leftover opioids.

## ACKNOWLEDGEMENTS

**Acknowledgements:** We thank participating practitioners from the National Dental Practice-Based Research Network (PBRN); study coordinators at every site; Megan Moncur, MS, who provided suggestions to improve the study; and the study participants. Study coordinators were compensated through their employment and participants were compensated for participation. Ms Moncur and the National Dental PBRN practitioners were not compensated for their work.

## Human Ethics and Consent to Participate

The APP study protocol (NCT04509115) was approved by the institutional review board at each participating site. All participants provided written informed consent before enrollment.

## Funding

This study was supported by the Food and Drug Administration (FDA) of the US Department of Health and Human Services (HHS) as part of a financial assistance award [U01FD005938] totaling $6,119,692 with 100 percent funded by FDA/HHS. The contents are those of the author(s) and do not necessarily represent the official views of, nor an endorsement, by FDA/HHS, or the US Government. National Dental PBRN infrastructure funding that enabled dentist recruitment was provided by grant U19DE28717 from the NIH. The FDA collaborated on all key aspects of the study, including design and conduct of the study; collection, management, analysis, and interpretation of the data; preparation, review, or approval of the manuscript; and decision to submit the manuscript for publication and is represented by 7 authors of the article.

### Authors Contributions

Erfan Taherifard: conceptualization, formal analysis, writing original draft, writing review and editing.

Mitra Ahadpour: conceptualization, investigation, funding acquisition, resources, supervision, writing review and editing.

Fernanda Bellolio: investigation, resources, writing review and editing.

Nancy Chang: conceptualization, investigation, resources, writing review and editing.

Summer Allen: conceptualization, investigation, resources, writing review and editing.

Marie-Carmelle Elie-Turenne: investigation, resources, supervision, writing review and editing.

W. Jonathan Fillmore: investigation, resources, writing review and editing.

Gregg H. Gilbert: investigation, resources, writing review and editing. Rebekah Heckmann: investigation, resources, writing review and editing.

Patricia Koussis: conceptualization, funding acquisition, resources, writing review and editing.

Christine Lee: conceptualization, investigation, resources, writing review and editing.

Heather Lipkind: conceptualization, investigation, resources, supervision, writing review and editing.

Tamra E. Meyer: conceptualization, investigation, resources, supervision, writing review and editing.

Teryl K. Nuckols: investigation, resources, supervision, writing review and editing.

Jessica D. Ritchie: conceptualization, investigation, resources, writing review and editing.

Michael A. Pacanowski: conceptualization, writing review and editing.

David B. Page: investigation, resources, writing review and editing.

Mat Soukup: conceptualization, investigation, writing review and editing.

Stephen Tamang: conceptualization, investigation, resources, writing review and editing.

Sam Torbati: investigation, resources, supervision, writing review and editing.

Doug Wallace: investigation, resources, writing review and editing.

Lauren Walter: investigation, resources, writing review and editing.

Yueqin Zhao: conceptualization, investigation, writing review and editing.

Nilay D. Shah: conceptualization, funding acquisition, resources, writing review and editing.

Joseph S. Ross: conceptualization, investigation, funding acquisition, supervision, writing review and editing.

Molly Moore Jeffery: conceptualization, investigation, formal analysis, funding acquisition, resources, supervision, writing review and editing.

### Data Availability

The datasets generated and analyzed during the current study are not publicly available.

## REFERENCES

1. Dowell D, Ragan KR, Jones CM, Baldwin GT, Chou R. CDC Clinical Practice Guideline for Prescribing Opioids for Pain - United States, 2022. MMWR Recomm Rep. Nov 4 2022;71(3):1–95. doi:10.15585/mmwr.rr7103a1

2. Phillips JK, Ford MA, Bonnie RJ, et al. Pain management and the intersection of pain and opioid use disorder. Pain management and the opioid epidemic: Balancing societal and individual benefits and risks of prescription opioid use. National Academies Press (US); 2017.

3. Daoust R, Paquet J, Cournoyer A, et al. Side effects from opioids used for acute pain after emergency department discharge. Am J Emerg Med. Apr 2020;38(4):695–701. doi:10.1016/j.ajem.2019.06.001

4. Carrasco-Labra A, Polk DE, Urquhart O, et al. Evidence-based clinical practice guideline for the pharmacologic management of acute dental pain in adolescents, adults, and older adults: A report from the American Dental Association Science and Research Institute, the University of Pittsburgh, and the University of Pennsylvania. J Am Dent Assoc. Feb 2024;155(2):102–117.e9. doi:10.1016/j.adaj.2023.10.009

5. Nafziger AN, Barkin RL. Opioid Therapy in Acute and Chronic Pain. J Clin Pharmacol. Sep 2018;58(9):1111–1122. doi:10.1002/jcph.1276

6. National Center for Health Statistics. U.S. Overdose Deaths Decrease for Third Consecutive Year in 2025. August 31, 2026, https://www.cdc.gov/nchs/pressroom/releases/20260513.html

7. Substance Abuse and Mental Health Services Administration. Key Substance Use and Mental Health Indicators in the United States: Results from the 2025 National Survey on Drug Use and Health. 2026. HHS Publication No. PEP26-07-010, NSDUH Series H-61. Accessed August 31, 2025. https://www.samhsa.gov/data/sites/default/files/reports/rpt57150/2025-nsduh-annual-national-report.pdf

8. Lipari RN, Hughes A. How People Obtain the Prescription Pain Relievers They Misuse. The CBHSQ Report. Substance Abuse and Mental Health Services Administration (US); 2013:1–7.

9. Substance Abuse Mental Health Services Administration. Key substance use and mental health indicators in the United States: Results from the 2019 National Survey on Drug Use and Health. HHS Publication No PEP20-07-01-001, NSDUH Series H-55. 2020;

10. Rosen PE, Ramdin C, Leonard J, Ruck BE, Nelson LS, Calello DP. Trends in opioid exposures among young children reported to United States poison centers from 2016 to 2023. Clin Toxicol (Phila). Nov 2024;62(11):762–769. doi:10.1080/15563650.2024.2401598

11. Bicket MC, Long JJ, Pronovost PJ, Alexander GC, Wu CL. Prescription Opioid Analgesics Commonly Unused After Surgery: A Systematic Review. JAMA Surg. Nov 1 2017;152(11):1066–1071. doi:10.1001/jamasurg.2017.0831

12. Mallama CA, Greene C, Alexandridis AA, McAninch JK, Dal Pan G, Meyer T. Patient-Reported Opioid Analgesic Use After Discharge from Surgical Procedures: A Systematic Review. Pain Med. Jan 3 2022;23(1):29–44. doi:10.1093/pm/pnab244

13. Collins CL, England K, Conrad SW, et al. Patient-reported duration of opioid analgesic use after discharge from surgical procedures or other types of acute pain: a scoping review. Pain Med. Sep 1 2025;26(9):503–514. doi:10.1093/pm/pnaf029

14. US Food and Drug Administration. FDA Approves REMS Modification Advancing New Drug Disposal Option. Accessed October 8, 2026, https://www.fda.gov/drugs/drug-alerts-and-statements/fda-approves-rems-modification-advancing-new-drug-disposal-option

15. US Food and Drug Administration. Where and How to Dispose of Unused Medicines. Accessed July 2, 2026, https://www.fda.gov/consumers/consumer-updates/where-and-how-dispose-unused-medicines

16. Lawrence AE, Carsel AJ, Leonhart KL, et al. Effect of Drug Disposal Bag Provision on Proper Disposal of Unused Opioids by Families of Pediatric Surgical Patients: A Randomized Clinical Trial. JAMA Pediatr. Aug 1 2019;173(8):e191695. doi:10.1001/jamapediatrics.2019.1695

17. Jeffery MM, Bellolio F, Chang N, et al. Opioid Use and Pain Resolution for Acute Pain Among Opioid-Naive Patients. JAMA Netw Open. Jul 1 2026;9(7):e2621875. doi:10.1001/jamanetworkopen.2026.21875

18. Jeffery MM, Ahadpour M, Allen S, et al. Acute pain pathways: protocol for a prospective cohort study. BMJ Open. Jul 5 2022;12(7):e058782. doi:10.1136/bmjopen-2021-058782

19. Hughes TD, Nowak J, Sottung E, Mustafa A, Lingechetty G. Empowering Pharmacists: Strategies for Addressing the Opioid Crisis through a Public Health Lens. Pharmacy (Basel). May 23 2024;12(3)doi:10.3390/pharmacy12030082

20. Thakur T, Frey M, Chewning B. Communication between patients and health care professionals about opioid medications. Explor Res Clin Soc Pharm. Jun 2021;2:100030. doi:10.1016/j.rcsop.2021.100030

21. Gregorian R, Marrett E, Sivathanu V, et al. Safe Opioid Storage and Disposal: A Survey of Patient Beliefs and Practices. J Pain Res. 2020;13:987–995. doi:10.2147/jpr.S242825

22. Varisco TJ, Fleming ML, Bapat SS, Wanat MA, Thornton D. Health care practitioner counseling encourages disposal of unused opioid medications. J Am Pharm Assoc (2003). Nov–Dec 2019;59(6):809–815.e5. doi:10.1016/j.japh.2019.07.010

23. Wotman S, Demko CA, Victoroff K, Sudano JJ, Lalumandier JA. A multimethod investigation including direct observation of 3751 patient visits to 120 dental offices. Clin Cosmet Investig Dent. 2010;2:27–39.

24. Neprash HT, Everhart A, McAlpine D, Smith LB, Sheridan B, Cross DA. Measuring Primary Care Exam Length Using Electronic Health Record Data. Med Care. Jan 2021;59(1):62–66. doi:10.1097/mlr.0000000000001450

25. Tai-Seale M, McGuire TG, Zhang W. Time allocation in primary care office visits. Health Serv Res. Oct 2007;42(5):1871–94. doi:10.1111/j.1475-6773.2006.00689.x

26. Arwi GA, Schug SA. Potential for Harm Associated with Discharge Opioids After Hospital Stay: A Systematic Review. Drugs. Apr 2020;80(6):573–585. doi:10.1007/s40265-020-01294-z

27. Hudgins JD, Porter JJ, Monuteaux MC, Bourgeois FT. Prescription opioid use and misuse among adolescents and young adults in the United States: A national survey study. PLoS Med. Nov 2019;16(11):e1002922. doi:10.1371/journal.pmed.1002922

28. Neill LA, Kim HS, Cameron KA, et al. Who Is Keeping Their Unused Opioids and Why? Pain Med. Jan 1 2020;21(1):84–91. doi:10.1093/pm/pnz025

29. Yeung C, Novak CB, Antflek D, Baltzer HL. Opioid Medication Disposal Among Patients Following Hand Surgery. Hand (N Y). Jun 2023;18(4):701–707. doi:10.1177/15589447211063585

