## Supplement for "Clinician Counseling and Patient Storage and Disposal Practices for Opioids Prescribed for Acute Pain: A Multicenter Prospective Cohort Study"

Table of Contents

Supplementary Table 1 Characteristics of Participants Prescribed Opioids for Acute Pain by Clinical Setting	2

Supplementary Table 2 Patient-Reported Instructions Received on When to Take and Stop Opioids, Overall and by Clinical Setting	4

Supplementary Table 3 Adjusted Models for Receipt of Counseling on Opioid Disposal Among Opioid-Naïve Participants Prescribed Opioids for Acute Pain	5

Supplementary Table 4 Storage and Disposal Practices of Opioid-Naïve Patients Prescribed Opioids for Acute Pain, Stratified by Presence of Children in the Household	6

Supplementary Table 5 Adjusted Models for Opioid Storage and Leftover Opioid Practices Among Opioid-Naïve Participants Prescribed Opioids for Acute Pain	8

Supplementary Table 6 Post Hoc Assessment of the Impact of Missing Outcome Data on the Proportion of Participant with Leftover Prescribed Opioids	9

**Supplementary Table 1.** Characteristics of Participants Prescribed Opioids for Acute Pain by Clinical Setting

|  | Total,  No. (%) | Emergency departments,  No. (%) | Primary care or urgent care clinics,  No. (%) | Inpatient settings,  No. (%) | Dental practice,  No. (%) | P value |
| --- | --- | --- | --- | --- | --- | --- |
| All participants | 1708 (100) | 915 (53.6) | 307 (18.0) | 223 (13.1) | 263 (15.4) |  |
| Age group | | | | | | |
| 21 and under | 170 (10.0) | 65 (7.1) | 6 (2.0) | 9 (4.0) | 90 (34.2) | <0.001 |
| 22-34 | 551 (32.3) | 277 (30.3) | 75 (24.4) | 107 (48.0) | 92 (35.0) |  |
| 35-44 | 368 (21.6) | 212 (23.2) | 71 (23.1) | 59 (26.5) | 26 (9.9) |  |
| 45-54 | 267 (15.6) | 168 (18.4) | 59 (19.2) | 13 (5.8) | 27 (10.3) |  |
| 55-64 | 216 (12.7) | 134 (14.6) | 48 (15.6) | 16 (7.2) | 18 (6.8) |  |
| 65 and older | 136 (8.0) | 59 (6.5) | 48 (15.6) | 19 (8.5) | 10 (3.8) |  |
| Children in household | | | | | | |
| Yes | 814 (71.2) | 424 (70.4) | 162 (67.5) | 103 (83.7) | 125 (69.8) | 0.01 |
| No | 330 (28.8) | 178 (29.6) | 78 (32.5) | 20 (16.3) | 54 (30.2) |  |
| Did not respond to survey | 564 | 313 | 67 | 100 | 84 | — |
| Race/ethnicity | | | | | | |
| White | 1093 (64.0) | 575 (62.8) | 268 (87.3) | 90 (40.4) | 160 (60.8) | <0.001 |
| Black or African American | 298 (17.5) | 142 (15.5) | 9 (2.9) | 79 (35.4) | 68 (25.9) |  |
| Hispanic or Latino | 155 (9.1) | 92 (10.1) | 11 (3.6) | 39 (17.5) | 13 (4.9) |  |
| American Indian or Alaska Native | 64 (3.8) | 45 (4.9) | 8 (2.6) | 11 (4.9) | 0 (0.0) |  |
| Asian | 43 (2.5) | 22 (2.4) | 3 (1.0) | 3 (1.4) | 15 (5.7) |  |
| Native Hawaiian or other Pacific Islander | 7 (0.4) | 5 (0.6) | 1 (0.3) | 0 (0.0) | 1 (0.4) |  |
| Other | 48 (2.8) | 34 (3.7) | 7 (2.3) | 1 (0.5) | 6 (2.3) |  |
| Residential setting | | | | | | |
| Urban | 647 (80.8) | 385 (87.3) | 135 (61.9) | 94 (93.1) | 33 (80.5) | <0.001 |
| Rural | 154 (19.2) | 56 (12.7) | 83 (38.1) | 7 (6.9) | 8 (19.5) |  |
| Did not answer | 353 | 279 | 57 | 69 | 149 | — |
| Did not respond to survey | 554 | 195 | 32 | 53 | 73 | — |
| Educational level | | | | | | |
| Less than high school | 104 (7.7) | 38 (5.3) | 10 (3.6) | 10 (6.5) | 46 (22.9) | <0.001 |
| High school diploma or GED | 342 (25.3) | 180 (25.0) | 66 (23.7) | 39 (25.3) | 57 (28.4) |  |
| More than high school | 906 (67.0) | 501 (69.7) | 202 (72.7) | 105 (68.2) | 98 (48.8) |  |
| Did not answer | 72 | 46 | 1 | 17 | 8 | — |
| Did not respond to survey | 284 | 150 | 28 | 52 | 54 | — |
| Pain source | | | | | | |
| Dental | 302 (17.7) | 27 (3.0) | 12 (3.9) | 0 (0.0) | 263 (100.0) | <0.001 |
| Obstetric | 176 (10.3) | 3 (0.3) | 1 (0.3) | 172 (77.1) | 0 (0.0) |  |
| Surgical or procedural | 65 (3.8) | 9 (1.0) | 5 (1.6) | 51 (22.9) | 0 (0.0) |  |
| Trauma or injury | 302 (17.7) | 219 (23.9) | 83 (27.0) | 0 (0.0) | 0 (0.0) |  |
| Musculoskeletal | 131 (7.7) | 70 (7.7) | 61 (19.9) | 0 (0.0) | 0 (0.0) |  |
| Abdominal | 98 (5.7) | 89 (9.7) | 9 (2.9) | 0 (0.0) | 0 (0.0) |  |
| Kidney stone | 88 (5.2) | 78 (8.5) | 10 (3.3) | 0 (0.0) | 0 (0.0) |  |
| Skin or soft tissue | 85 (5.0) | 61 (6.7) | 24 (7.8) | 0 (0.0) | 0 (0.0) |  |
| Low back pain | 100 (5.9) | 61 (6.7) | 39 (12.7) | 0 (0.0) | 0 (0.0) |  |
| Other | 86 (5.0) | 65 (7.1) | 21 (6.8) | 0 (0.0) | 0 (0.0) |  |
| Unknown in emergency department | 233 (13.6) | 233 (25.5) | 0 (0.0) | 0 (0.0) | 0 (0.0) |  |
| Unknown in primary Care or urgent care clinics | 42 (2.5) | 0 (0.0) | 42 (13.7) | 0 (0.0) | 0 (0.0) |  |

**Note:** Percentages are calculated among participants with nonmissing responses for each characteristic. “Did not answer” indicates participants who responded to some questions on the relevant survey but did not respond to that item. “Did not respond to survey” indicates participants who did not complete the relevant survey.

**Abbreviation:** GED, General Educational Development.

**Supplementary Table 2.** Patient-Reported Instructions Received on When to Take and Stop Opioids, Overall and by Clinical Setting

| Instructions received | Overall, No. (%) | Emergency departments, No. (%) | Primary care or urgent care clinics, No. (%) | Inpatient settings, No. (%) | Dental practice, No. (%) | P value |
| --- | --- | --- | --- | --- | --- | --- |
| When to take opioids | | | | | | |
| All participants with available data | 1328 (100) | 713 (100) | 277 (100) | 146 (100) | 192 (100) | — |
| Take opioids only when pain is very bad | 522 (39.3) | 308 (43.2) | 115 (41.5) | 36 (24.7) | 63 (32.8) | <0.001 |
| Take opioids at the first sign of pain | 317 (23.9) | 180 (25.2) | 63 (22.7) | 30 (20.5) | 44 (22.9) | 0.59 |
| Take opioids on a regular schedule, whether or not there is pain | 193 (14.5) | 97 (13.6) | 28 (10.1) | 34 (23.3) | 34 (17.7) | 0.001 |
| Take opioids at specific times, whether or not there is pain | 110 (8.3) | 49 (6.9) | 27 (9.7) | 17 (11.6) | 17 (8.9) | 0.18 |
| Try other medications first and take opioids only if they do not work | 291 (21.9) | 137 (19.2) | 64 (23.1) | 26 (17.8) | 64 (33.3) | <0.001 |
| No instructions on when to take opioids | 101 (7.6) | 48 (6.7) | 20 (7.2) | 26 (17.8) | 7 (3.6) | <0.001 |
| When to stop opioids | | | | | | |
| All participants with available data | 1302 (100) | 694 (100) | 275 (100) | 138 (100) | 195 (100) | — |
| Stop after a specified number of days | 177 (13.6) | 85 (12.2) | 33 (12.0) | 26 (18.8) | 33 (16.9) | 0.08 |
| Stop when the pills run out | 289 (22.2) | 168 (24.2) | 57 (20.7) | 26 (18.8) | 38 (19.5) | 0.30 |
| Continue taking opioids as long as there is pain | 417 (32.0) | 212 (30.5) | 88 (32.0) | 36 (26.1) | 81 (41.5) | 0.01 |
| No instructions on when to stop opioids | 456 (35.0) | 250 (36.0) | 98 (35.6) | 54 (39.1) | 54 (27.7) | 0.11 |

**Note:** Participants could report more than one reason.

Supplementary Table **3.** Adjusted Models for Receipt of Counseling on Opioid Disposal Among Opioid-Naïve Participants Prescribed Opioids for Acute Pain

| Characteristic | Counseling on importance of opioid disposal,  RR (95% CI) | P value | Counseling on appropriate disposal methods,  RR (95% CI) | P value |
| --- | --- | --- | --- | --- |
| Age group | | | | |
| ≤21 | Reference | — | Reference | — |
| 22-34 | 1.01 (0.76-1.34) | 0.97 | 0.92 (0.68-1.25) | 0.60 |
| 35-44 | 0.95 (0.69-1.29) | 0.72 | 0.93 (0.67-1.28) | 0.64 |
| 45-54 | 0.88 (0.63-1.23) | 0.45 | 0.87 (0.62-1.23) | 0.44 |
| 55-64 | 0.89 (0.63-1.26) | 0.50 | 0.96 (0.67-1.36) | 0.81 |
| ≥65 | 0.95 (0.64-1.42) | 0.82 | 0.90 (0.59-1.37) | 0.61 |
| Clinical setting | | | | |
| Emergency departments | Reference | — | Reference | — |
| Dental practice | 0.99 (0.77-1.27) | 0.93 | 1.07 (0.82-1.38) | 0.63 |
| Inpatient settings | 1.02 (0.79-1.31) | 0.89 | 1.12 (0.86-1.46) | 0.40 |
| Primary care or urgent care clinics | 0.94 (0.77-1.16) | 0.57 | 0.99 (0.79-1.23) | 0.91 |
| Children in household | | | | |
| No | Reference | — | Reference | — |
| Yes | 1.00 (0.83-1.20) | 0.96 | 1.01 (0.83-1.24) | 0.90 |
| Missing response | 1.19 (0.90-1.57) | 0.22 | 1.24 (0.93-1.67) | 0.15 |

**Note:** Risk ratios were calculated as ratios of predictive margins after logistic regression with robust standard errors. Models included age group, clinical setting, and presence of children in the household. Receipt of counseling was defined as reporting that counseling was provided by the prescriber, pharmacist, or both. Abbreviation: CI, confidence interval; RR, risk ratio.

Supplementary Table **4.** Storage and Disposal Practices of Opioid-Naïve Patients Prescribed Opioids for Acute Pain, Stratified by Presence of Children in the Household

|  | Patients Provided Information on Household Children No. (%) | No Children in Household  No. (%) | Children in Household  No. (%) | P Value |
| --- | --- | --- | --- | --- |
| All patients | 1144 (100) | 330 (100) | 814 (100) | — |
| Storing opioids in a latched location | | | | |
| Never | 292 (39.7) | 110 (48.0) | 182 (35.9) | 0.006 |
| Almost never | 48 (6.5) | 16 (7.0) | 32 (6.3) |  |
| Sometimes | 37 (5.0) | 11 (4.8) | 26 (5.1) |  |
| Almost always | 36 (4.9) | 14 (6.1) | 22 (4.3) |  |
| Always | 323 (43.9) | 78 (34.1) | 245 (48.3) |  |
| Did not use opioid | 292 | 65 | 227 | — |
| Did not answer | 116 | 36 | 80 | — |
| Storing opioids in a locked location | | | | |
| Never | 523 (76.1) | 178 (83.2) | 345 (72.9) | 0.02 |
| Almost never | 36 (5.2) | 11 (5.1) | 25 (5.3) |  |
| Sometimes | 15 (2.2) | 3 (1.4) | 12 (2.5) |  |
| Always | 113 (16.5) | 22 (10.3) | 91 (19.2) |  |
| Did not use opioid | 13 | 4 | 9 | — |
| Did not answer | 444 | 112 | 332 | — |
| Opioid storage location | | | | |
| Original prescription bottle | | | | |
| Yes | 682 (66.4) | 215 (73.1) | 467 (63.6) | 0.004 |
| No | 346 (33.6) | 79 (26.9) | 267 (36.4) |  |
| Did not answer | 116 | 36 | 80 | — |
| Daily or weekly pill organizer | | | | |
| Yes | 9 (0.9) | 2 (0.7) | 7 (1.0) | 0.67 |
| No | 1019 (99.1) | 292 (99.3) | 727 (99.0) |  |
| Did not answer | 116 | 36 | 80 | — |
| Single-compartment container | | | | |
| Yes | 12 (1.2) | 2 (0.7) | 10 (1.4) | 0.36 |
| No | 1016 (98.8) | 292 (99.3) | 724 (98.6) |  |
| Did not answer | 116 | 36 | 80 | — |
| Use of all prescribed opioids | | | | |
| Yes | 258 (33.4) | 72 (30.8) | 186 (34.5) | 0.31 |
| No | 515 (66.6) | 162 (69.2) | 353 (65.5) |  |
| Did not answer | 371 | 96 | 275 | — |
| Kept leftover opioids among those who did not use all opioids and provided information on household children (n=515) | | | | |
| Yes | 214 (42.5) | 84 (53.2) | 130 (37.7) | 0.001 |
| No | 289 (57.5) | 74 (46.8) | 215 (62.3) |  |
| Did not answer | 12 | 4 | 8 | — |
| Disposal methods used among those who disposed of leftover opioids and provided information on household children (n=289)^a^ | | | | |
| Return to take-back location | | | | |
| Yes | 99 (41.0) | 25 (40.3) | 74 (41.1) | 0.91 |
| No | 143 (59.0) | 37 (59.7) | 106 (58.9) |  |
| Did not answer | 47 | 12 | 35 | — |
| Flush down the toilet | | | | |
| Yes | 91 (37.6) | 26 (41.9) | 65 (36.1) | 0.43 |
| No | 151 (62.4) | 36 (58.1) | 115 (63.9) |  |
| Did not answer | 47 | 12 | 35 | — |
| Throw away in the trash | | | | |
| Yes | 55 (22.7) | 12 (19.4) | 43 (23.9) | 0.46 |
| No | 187 (77.3) | 50 (80.6) | 137 (76.1) |  |
| Did not answer | 47 | 12 | 35 | — |

**Note:** Percentages for storage-related items are calculated among participants with nonmissing responses who reported opioid use. “Did not answer” indicates participants who responded to some questions on the relevant survey but did not respond to that item. “Did not use opioid” indicates participants who reported no opioid use at the time of survey completion. Participants may have reported more than one disposal method.

Supplementary Table **5.** Adjusted Models for Opioid Storage and Leftover Opioid Practices Among Opioid-Naïve Participants Prescribed Opioids for Acute Pain

| Characteristic | Storing opioids in a latched or locked location,  RR (95% CI) | P value | Kept leftover opioids among participants who did not use all prescribed opioids,  RR (95% CI) | P value |
| --- | --- | --- | --- | --- |
| Age group | | | | |
| ≤21 | Reference | — | Reference | — |
| 22-34 | 1.29 (0.95-1.76) | 0.11 | 1.03 (0.94-1.14) | 0.48 |
| 35-44 | 1.35 (0.98-1.87) | 0.06 | 1.05 (0.95-1.16) | 0.32 |
| 45-54 | 1.26 (0.91-1.75) | 0.17 | 1.05 (0.96-1.16) | 0.29 |
| 55-64 | 1.10 (0.77-1.56) | 0.61 | 1.05 (0.96-1.16) | 0.28 |
| ≥65 | 0.82 (0.53-1.28) | 0.39 | 1.01 (0.88-1.15) | 0.90 |
| Clinical setting | | | | |
| Emergency departments | Reference | — | Reference | — |
| Dental practice | 0.97 (0.79-1.20) | 0.78 | 1.02 (0.99-1.06) | 0.15 |
| Inpatient settings | 1.04 (0.85-1.28) | 0.71 | 1.01 (0.96-1.06) | 0.84 |
| Primary care or urgent care clinics | 0.89 (0.75-1.07) | 0.22 | 1.02 (0.99-1.05) | 0.28 |
| Children in household | | | | |
| No | Reference | — | Reference | — |
| Yes | 1.25 (1.05-1.48) | 0.01 | 1.00 (0.97-1.03) | 1.00 |
| Missing response | 1.12 (0.89-1.40) | 0.33 | 1.00 (0.96-1.03) | 0.82 |

**Note:** Risk ratios were calculated as ratios of predictive margins after logistic regression with robust standard errors. Storing opioids in a latched or locked location was defined as responding always or almost always to how often participants stored opioids in a latched or locked location. Models included age group, clinical setting, and presence of children in the household. The participants included in each model varied according to the outcome assessed and the availability of relevant data. Abbreviation: CI, confidence interval; RR, risk ratio.

Supplementary Table **6.** Post Hoc Assessment of the Impact of Missing Outcome Data on the Proportion of Participant with Leftover Prescribed Opioids

| Analysis | Participants with leftover opioids, % (95% CI) |
| --- | --- |
| Complete-case analysis (n=929) | 66.5 (63.4-69.5) |
| Multiple imputation analysis (n=1708) | 65.1 (61.4-69.0) |

**Note:** Data on whether participants had leftover prescribed opioids were missing for 779 of 1708 participants (45.6%). A post hoc multiple imputation analysis was conducted to assess the potential impact of missing outcome data. Missing outcome values were imputed using logistic regression with age group, clinical setting, presence of children in the household, race/ethnicity, education level, and pain source as predictors. Missing predictor values were included in the analysis as a separate missing category (e.g., presence of children in the household was categorized as yes, no, or missing). A total of 50 imputations were completed and analyzed using Stata version 19.0 mi commands.
